# Open Science and COVID-19 Randomized Controlled Trials: Examining Open Access, Preprinting, and Data Sharing-Related Practices During the Pandemic

**DOI:** 10.1101/2022.08.10.22278643

**Authors:** John A. Borghi, Cheyenne Payne, Lily Ren, Amanda L. Woodward, Connie Wong, Christopher Stave

## Abstract

The COVID-19 pandemic has brought substantial attention to the systems used to communicate biomedical research. In particular, the need to rapidly and credibly communicate research findings has led many stakeholders to encourage researchers to adopt open science practices such as posting preprints and sharing data. To examine the degree to which this has led to the adoption of such practices, we examined the “openness” of a sample of 539 published papers describing the results of randomized controlled trials testing interventions to prevent or treat COVID-19. The majority (56%) of the papers in this sample were free to read at the time of our investigation and 23.56% were preceded by preprints. However, there is no guarantee that the papers without an open license will be available without a subscription in the future, and only 49.61% of the preprints we identified were linked to the subsequent peer-reviewed version. Of the 331 papers in our sample with statements identifying if (and how) related datasets were available, only a paucity indicated that data was available in a repository that facilitates rapid verification and reuse. Our results demonstrate that, while progress has been made, there is still a significant mismatch between aspiration and the practice of open science in an important area of the COVID-19 literature.

**Open Materials:** We are committed to making the details of our research process as open as possible. The data and code that underlie our analyses are archived and published through the Dryad Data Repository (https://doi.org/10.5061/dryad.mkkwh7137). Documentation and instructions for manuscript screening and data extraction are available on Protocols.io (https://dx.doi.org/10.17504/protocols.io.x54v9jx7zg3e/v1). Author contributions are outlined in Supplementary Table 1.

## Introduction

Over 200,000 COVID-19-related preprints and publications were published between January 2020 and January 2022^1^ and the characteristics of this immense body of work have catalyzed vital conversations about how research is conducted, evaluated, and communicated. Since the onset of the pandemic, funding agencies, scholarly publishers and other stakeholders have worked to ensure that COVID-19-related research is rapidly and broadly disseminated (Waltman et al., 2021). In January 2020, over 160 research funders, scholarly publishers, and other organizations signed a statement issued by the Wellcome Trust that committed signatories to work together to ensure that peer-reviewed publications related to COVID-19 are openly or freely accessible, that related research findings are disseminated rapidly through preprint servers prior to their publication, and that research data and other materials are shared as quickly and broadly as possible (Wellcome Trust, 2020). In March 2020, the science advisors from twelve countries urged scholarly publishers to voluntarily agree to make their COVID-19 and coronavirus-related publications and datasets immediately accessible in PubMed Central and other public repositories (WHOSTP, 2020).

These activities - ensuring access to publications, posting preprints, and data sharing - fall under the umbrella of “open science”, which covers a wide variety of efforts focused on making scientific research more accessible and transparent (NASEM, 2018). Practices related to open science exist along a continuum where individual research efforts can be described as more or less “open” along a number of dimensions. For certain types of research outputs, such as datasets, ensuring accessibility does not necessarily mean ensuring availability to everyone for any purpose. “Openness” in this context could be interpreted as ensuring there is a clearly defined pathway for requesting and, when appropriate, being granted access.

In biomedicine and the health sciences, open science practices have been put forward as methods for both increasing the speed and credibility of the research process and also catalyzing innovation (Chan et al., 2014). Especially notable in the context of COVID-19 has been the unprecedented dissemination of preprints - early versions of articles that have not yet gone through the peer review process - to rapidly communicate research findings (Fraser et al., 2021)

For randomized controlled trials (RCTs), which are frequently described as representing the “gold standard” of evidence (Jones & Podolsky, 2015), the importance of establishing the transparency, reproducibility, and validity of results is paramount. To assess the degree to which the converging statements, policies, and initiatives designed to facilitate the communication of research results related to COVID-19 have resulted in the adoption of open science practices, we examined the openness of publications detailing the results of RCTs for pharmaceutical interventions designed to prevent or treat the virus. For this study, we define “openness” quite broadly, to include the accessibility of the publications themselves, if they were preceded by preprints, and if they provided information about how to obtain related datasets and other materials.

## Methods

To locate published reports of COVID-19-related clinical trials, we conducted a comprehensive literature search for publications describing the results of RCTs in PubMed, Embase, and the Cochrane Central Register of Clinical Trials. The search strings for each database are available in Supplementary Table 2. To be included in our dataset, a publication had to describe the results of a randomized trial for a pharmaceutical intervention to prevent or treat COVID-19. Meeting abstracts, posters, commentaries, preprints, retracted publications, and publications describing non-randomized trials or trials with non-human participants were excluded as were publications examining RCTs for behavioral interventions (e.g. use of personal protective equipment). The publications in our sample were published prior to January 1st, 2022. Each paper was screened by at least two members of the research team. To examine the relationship between the adoption of open science practices and the communication of scientific results, we extracted citation counts and Altmetric scores for each paper using Dimensions (https://app.dimensions.ai/).

Since 2004, the International Committee of Medical Journal Editors (ICMJE) has recommended that all medical journal editors require the registration of clinical trials in a public trials registry as a condition of consideration for publication (De Angelis et al., 2004). Following the completion of the screening process, we manually extracted information about how and where the trials described by each paper were registered and what interventions were tested.

We identified the accessibility of the publications in our sample using two data sources. In light of the Public Health Emergency COVID-19 Initiative (National Library of Medicine, 2020), which has resulted in the deposit of COVID-19 related publications into PubMed Central, we first checked for copies of each paper in that repository. We then used the Unpaywall database (Piwowar et al., 2018) to retrieve more detailed information about the mechanisms by which each paper was being made open.

To identify whether a publication was preceded by a preprint, we searched Europe PMC (https://europepmc.org), a platform that indexes the majority of preprint platforms serving the biomedical and health sciences. If a title match was not located, we searched for preprints credited to the same authors through Europe PMC and Google Scholar. Information about the preprint policies of individual journals was extracted from Sherpa Romeo (https://v2.sherpa.ac.uk/romeo).

Since 2017, ICMJE member journals have required that papers describing the results of RCTs be accompanied by data availability statements that detail how to access related data and other materials (Taichman et al., 2017). When such statements were available, we examined whether or not they indicated that related datasets are or will be made available, through what mechanisms data was said to be available, and, for datasets noted as available through data repositories, what repositories were indicated and if a URL or persistent identifier was provided. Because of the ambiguity of some of the data availability statements in our sample, we coded statements as indicating data availability if they did not explicitly state that data was unavailable.

Data analysis and visualization were performed in R, using the dplyr (Wickham et al., 2022), ggplot (Wickham, 2016), and waffle (Rudis et al., 2019) packages. Unpaired two-sample t-tests were used to evaluate the impact of ICMJE membership, ICMJE recommendation, and preprint availability on citation counts and Altimetric scores.

## Results

After screening, we identified 539 publications that met our inclusion criteria. Of these, 106 (19.67%) were published in ICMJE member journals and 259 (48.05%) were published in journals that state that they follow ICMJE recommendations (e.g. requiring trial registration, inclusion of a data availability statements). The majority (n = 447, 83%) were published by a signatory of the statement issued by the Wellcome Trust. Papers from ICMJE member journals received significantly higher numbers of citations [*t*(104.35) = 5.318, p < 0.001] and Altmetric scores [*t*(106.79) = 6.387, p < 0.001] as did journals that state that they follow ICMJE recommendations (which includes ICMJE member journals) [*t*(275.35) = 4.732, p < 0.001; *t*(306.75) = 5.370, p < 0.001] when compared to the other papers in our sample.

We were able to locate registration information for the trials underlying 482 (89.42%) of the papers in our sample. Trials were most commonly registered via ClinicalTrials.gov (n = 319). The most common interventions in our sample were convalescent plasma (26 papers), hydroxychlorquine (22 papers), and ivermectin (22 papers).

We were able to locate a copy of the majority of the papers in PubMed Central (n = 487, 90.72%). As shown in Table 1, according to Unpaywall, the most common mechanism by which the papers in our sample were made available was via the publishing journal’s website (221 papers, 41.00%).

**Table 1.** Mechanism of availability of published papers describing results of randomized controlled trials for pharmaceutical interventions to prevent or treat COVID-19. Using the Unpaywall database, we were able to find information regarding the availability for 529 of the 539 papers in our sample. At the time of our investigation, only a small number were not openly accessible, though it is possible that articles made “free to read” without an explicit open license may require a subscription to access in the future.

| Mechanism of Paper Availability | Count |
| --- | --- |
| Made free to read on publishing journal's website | 221 (41.00%) |
| Published in an an open access journal | 198 (36.73%) |
| Openly available in a subscription-based journal | 65 (12.06%) |
| Available through an open access repository or archive | 28 (5.19%) |
| Available only via subscription (Closed) | 17 (3.21%) |

Of the 539 papers in our sample, 437 (81.08%) were published in a journal that does not prohibit the posting of a preprint on a public website or repository. We were able to locate preprints for 127 (23.56% of our total sample, 29.06% of those in journals with preprint policies) across 4 different preprint servers, the most common being MedRxiv (76 preprints). Of the preprints we were able to identify, 87 (68.50%) had a different title than the published version and only 63 (49.61%) included a direct link to the published version. In contrast to previous research indicating that preprints may confer a citation advantage (Fu & Hughey, 2019), no significant differences in either citation number [*t*(198.6) = 0.559, p = 0.579] or Altmetric score [*t*(345.58) = 0.412, p = 0.681] were found between the papers in our sample preceded and not preceded by preprints.

We were able to locate data availability statements for 331 (61.41%) of the papers in our sample, with 303 (56.21% of total papers, 91.54% of papers with statements) indicating that the data is available through some mechanism. As shown in figure 1. and table 2., the majority of these statements indicated that the data was available by request, either to an author or to another entity (e.g. the study sponsor). Only 19 of the papers with statements in our sample (5.74%) indicated that related datasets were available via a repository - with just 7 (2.11%) giving a URL or persistent identifier that resolved to a page where the data could be accessed.

**Figure 1.**
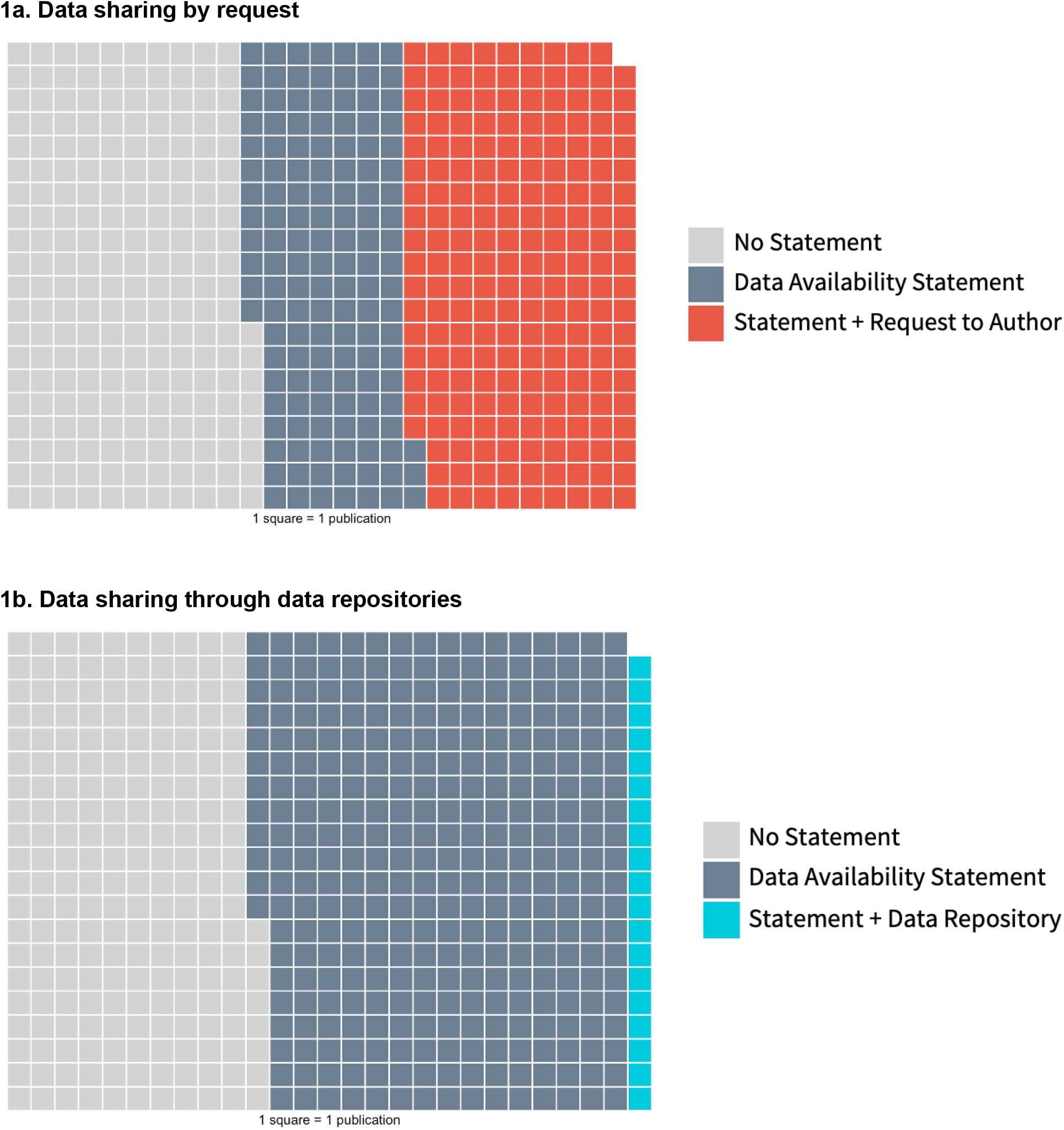
Methods of data sharing. Visual representation of data sharing in our sample. Each box represents one paper. 1A: The most common mechanism of making data available was via request, either to the author or to another entity. 1B: Of the 19 papers with data availability statements stating that data is available through a repository, only 12 statements included a link that resolved to the datasets and only 13 datasets could be found by using the link or through manually searching the repository.

**Table 2.** Mechanism of data availability for published papers describing results of randomized controlled trials for pharmaceutical interventions to prevent or treat COVID-19. By manually locating and extracting information from data availability statements we were able to gather information about the mechanisms through which the datasets underlying the results of the papers in our sample are available. Percentages are calculated out of number of papers with data availability statements (331). Other than checking data repositories, we did not attempt to validate any of these statements (e.g. by requesting data). Examining the reproducibility of the datasets available through any of these mechanisms was also beyond the scope of this study.

| Mechanism of Data Availability | Count |
| --- | --- |
| Request to author | 196 (59.21%) |
| Request to other (e.g. study sponsor) | 55 (16.62%) |
| Publication or supplementary materials | 25 (7.55%) |
| Data Repository | 19 (5.74%) |
| Trial Registry | 5 (1.51%) |
| Other | 3 (0.91%) |
| Unclear | 13 (3.93%) |

Overall, of the 539 papers in our sample, we were able to identify a total of 89 (16.51%) that were free to read, were preceded by a preprint, and carried a data availability statement that stated that data are available. Only 4 papers (0.7%) were free to read, preceded by a preprint that linked to the published version, and were accompanied by a statement that stated that data is available in a data repository (with only 3 providing a direct link to where the data could be accessed).

## Discussion

The ability to rapidly access, evaluate, and build upon research results has been essential to the worldwide response to COVID-19. Perhaps the clearest demonstration of this has been the sharing of the complete SARS-COV-2 genome in January 2020. Initially shared through the Virological discussion forum and subsequently through the Genbank repository (GenBank accession number: MN908947; see also Wu et al., 2020), the open publication of this sequence catalyzed the development of tests and treatments for the disease.

In this study, our goal was to examine a segment of the scholarly communications landscape - randomized controlled trials for pharmaceutical interventions intended to prevent or treat COVID-19 - to determine if a constellation of statements, policies, and initiatives has resulted in the adoption of open science practices. Our goal was not to address the content of these studies. Rather, we aimed to assess the extent to which published papers were openly available, preceded by preprints, or accompanied by data availability statements with directions to related datasets. We note that these attributes should not be interpreted as indicators of the rigor of the trials or the efficacy of the interventions being examined. Any appraisal of adherence to reporting guidelines (see Quinn et al., 2021) or evaluation of the strength of evidence (see Honarmand et al., 2021) for any of the individual studies in our sample or in the literature as a whole was beyond the scope of this study.

In line with work demonstrating the accessibility of COVID-19-related publications early in the pandemic (Arrizabalaga et al., 2020), our results show that a high percentage of the literature related to a particular topic can be made free to read using readily available mechanisms. However, our results also illustrate that these mechanisms and the papers made available through them exist along a continuum of openness. The most common mechanism by which the articles in our sample are available - through a website hosted by their publisher but potentially without a formal license for reuse - is suboptimal. The statement issued by the Wellcome Trust states that papers should be made free to read “for at least the duration of the outbreak” (Wellcome Trust, 2020). However, without a license ensuring reuse rights, access to these articles could be denied at any point in the future.

The dissemination of preprints in biomedical science dates back to the 1960’s (see Cobb, 2017), but the COVID-19 literature features preprints to an unprecedented degree (Fraser et al., 2021; Johansson et al., 2018). Preprints allow for rapid dissemination of vital research results, particularly for time-sensitive issues such as COVID-19. However, preprints may also be a source for questionable, fraudulent, or even just preliminary claims that can be taken up by individuals or outlets who are unaware that the source has not gone through the traditional peer-review process (Watson, 2022). Of course, questionable and fraudulent claims can also be present in peer reviewed literature ((Ledford & Van Noorden, 2020)and, while substantial changes *can* occur between a given paper’s preprint and peer-reviewed version, the majority of these changes do not appear to qualitatively affect the paper’s conclusion (Brierley et al., 2022).

Our results are indicative of a movement towards posting preprints in biomedical and health sciences research. To allay concern about the posting of preprints, we recommend that preprints be clearly marked as not yet peer-reviewed and linked to the final version when available. While the preprints preceding the articles in our sample were all available on servers that clearly marked them as not yet peer-reviewed, many were not linked to the final version, even when the preprint server and journal were operated by the same publisher.

Access to the datasets underlying the results described in preprints and published work is foundational to establishing their transparency, reproducibility, and validity (Ewers et al., 2021) and datasets from clinical trials is evidence synthesis. However, compared to peer-reviewed articles (and preprints), openness for data presents a number of complex ethical, legal, and technical challenges. Addressing these requires the implementation of a variety of technological solutions and governance structures (Mangravite et al., 2020). In line with previous analyses demonstrating a growing intent to share COVID-19 data (Larson et al., 2022; Li et al., 2021), we found that most of the data availability statements for the papers in our sample stated that the data was ostensibly available (or, at least, not explicitly unavailable). Unfortunately, the most common mechanism by which data was stated to be available - by request to an author or other party - has been shown to be substantially less than optimal. A number of studies (e.g. Gabelica et al., 2022) have demonstrated that data stated to be available in this manner are typically not in practice.

The widespread use of suboptimal data sharing methods and the ambiguity of many of the data availability statements in our sample points to the need to provide specific guidance to researchers about how to share data and what should be included in a data availability statement. If the data is available in a repository, even one with restricted access, a persistent identifier should be included to facilitate data discovery. If data cannot be made available through a repository, then the statement should include specific and reliable information about the mechanism for requesting access and how such requests will be evaluated and acted upon.

Taken together, our results reflect the changing scholarly communications landscape amid COVID-19. At least for RCTs examining interventions to prevent or treat COVID-19, progress has been made in terms of making papers, preprints, and datasets available, but there is still a significant mismatch between aspiration and actual practice.

Ensuring that scientific research is accessible and transparent is broader than just addressing the availability of discrete research outputs. In order to examine the “openness” of the papers in our sample, we needed to look across a variety of data sources. One major takeaway from our research process is that finding one output of a particular research effort (such as a dataset or preprint) even with access to another output of that same effort (such as the publication) is, in practice, not straightforward. The technical means for addressing this are already available. Researchers, papers, datasets, and other elements connected to a particular research effort can be easily and unambiguously linked using persistent identifiers (e.g. ORCID iDs, DOIs, RRIDs, etc). However, realizing the benefits of open science requires researchers, policymakers, and other stakeholders to implement strategies and practices throughout the entire research process - starting before data is collected and continuing well after related papers are published (Borghi & Van Gulick, 2022).

The COVID-19 pandemic has demonstrated the need for science to be rapidly and transparently communicated. To conclude we offer two recommendations for closing the gap between aspiration and the practice of open science for pandemic-related research.

1. Increased coordination between research stakeholders to establish and maintain links between accessible research outputs. This includes the use of persistent identifiers but could also mean exposing such links in a more human readable fashion, such as through an “open materials” or “related works” section on a journal, server, or repository page.
2. Increased guidance and assistance for researchers to facilitate the integration of open science practices into their research workflows. This includes, but is certainly not limited to specific instructions on methods for sharing data effectively (e.g. through a specialized data repository).

## Data Availability

We are committed to making the details of our research process as open as possible. The data and code that underlie our analyses are archived and published through the Dryad Data Repository (https://doi.org/10.5061/dryad.mkkwh7137). Documentation and instructions for manuscript screening and data extraction are available on Protocols.io (https://dx.doi.org/10.17504/protocols.io.x54v9jx7zg3e/v1). Author contributions are outlined in Supplementary Table 1.

---

**Supplementary Table 1.**
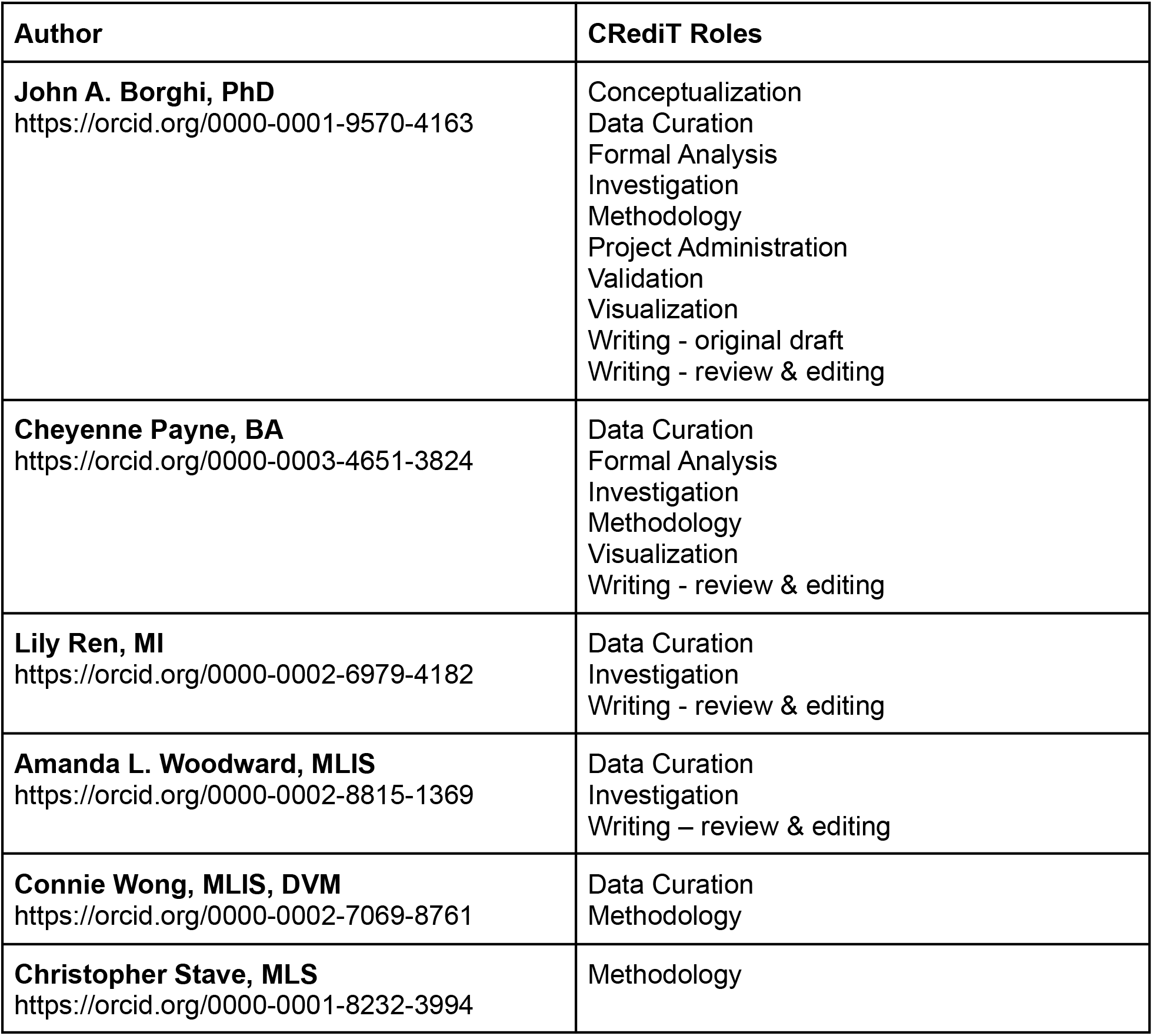
Author Information and Contributions.

**Supplementary Table 2.**
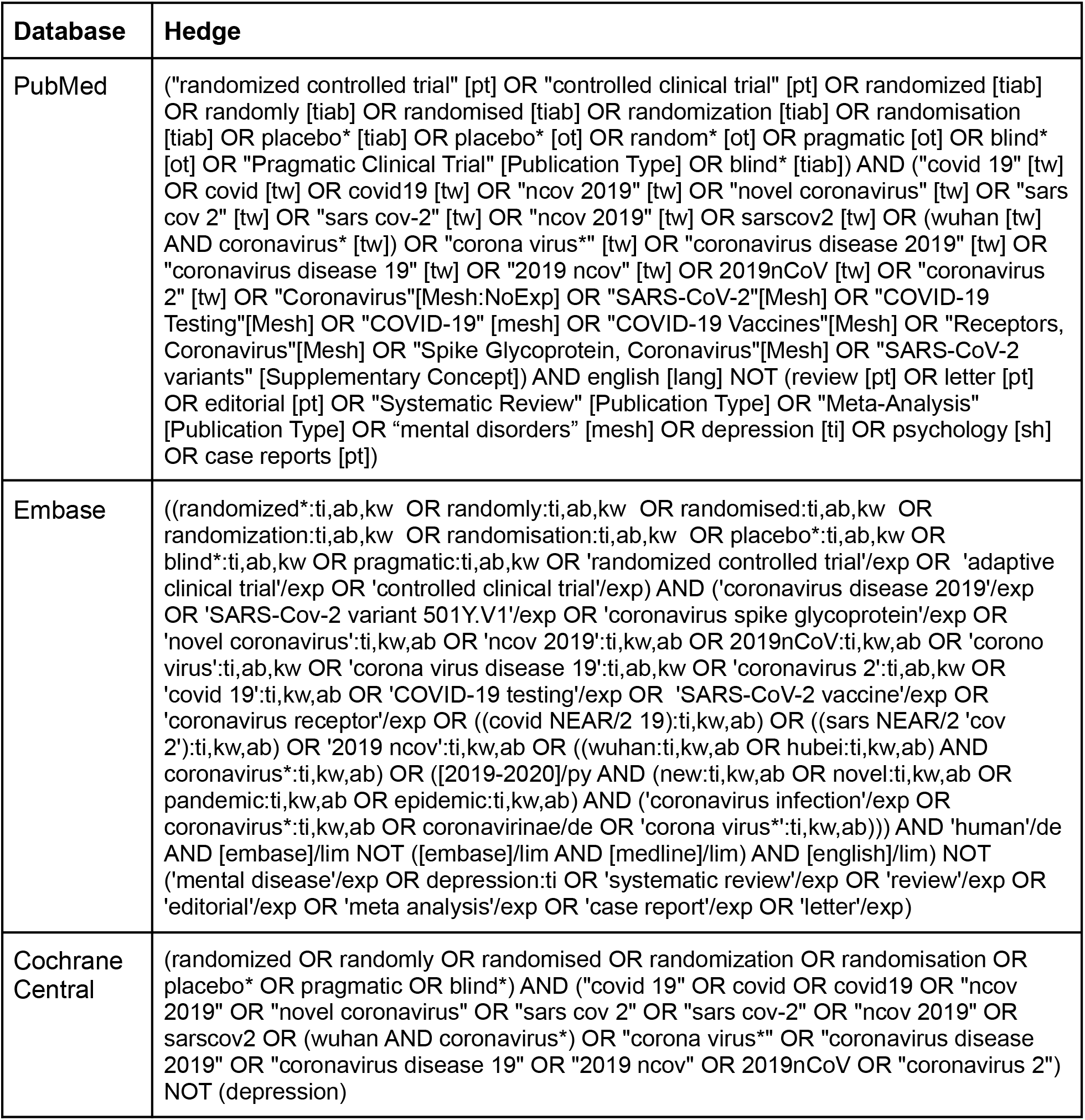
Search hedges for the three databases.

| Database | Hedge |
| --- | --- |
| PubMed | ("randomized controlled trial" [pt] OR "controlled clinical trial" [pt] OR randomized [tiab] OR randomly [tiab] OR randomised [tiab] OR randomization [tiab] OR randomisation [tiab] OR placebo* [tiab] OR placebo* [ot] OR random* [ot] OR pragmatic [ot] OR blind* [ot] OR "Pragmatic Clinical Trial" [Publication Type] OR blind* [tiab]) AND ("covid 19" [tw] OR covid [tw] OR covid19 [tw] OR "ncov 2019" [tw] OR "novel coronavirus" [tw] OR "sars cov 2" [tw] OR "sars cov-2" [tw] OR "ncov 2019" [tw] OR sarscov2 [tw] OR (wuhan [tw] AND coronavirus* [tw]) OR "corona virus*" [tw] OR "coronavirus disease 2019" [tw] OR "coronavirus disease 19" [tw] OR "2019 ncov" [tw] OR 2019nCoV [tw] OR "coronavirus 2" [tw] OR "Coronavirus"[Mesh:NoExp] OR "SARS-CoV-2"[Mesh] OR "COVID-19 Testing"[Mesh] OR "COVID-19" [mesh] OR "COVID-19 Vaccines"[Mesh] OR "Receptors, Coronavirus"[Mesh] OR "Spike Glycoprotein, Coronavirus"[Mesh] OR "SARS-CoV-2 variants" [Supplementary Concept]) AND english [lang] NOT (review [pt] OR letter [pt] OR editorial [pt] OR "Systematic Review" [Publication Type] OR "Meta-Analysis" [Publication Type] OR "mental disorders" [mesh] OR depression [ti] OR psychology [sh] OR case reports [pt]) |
| Embase | ((randomized*:ti,ab,kw OR randomly:ti,ab,kw OR randomised:ti,ab,kw OR randomization:ti,ab,kw OR randomisation:ti,ab,kw OR placebo*:ti,ab,kw OR blind*:ti,ab,kw OR pragmatic:ti,ab,kw OR 'randomized controlled trial'/exp OR 'adaptive clinical trial'/exp OR 'controlled clinical trial'/exp) AND ('coronavirus disease 2019'/exp OR 'SARS-Cov-2 variant 501Y.V1'/exp OR 'coronavirus spike glycoprotein'/exp OR 'novel coronavirus':ti,kw,ab OR 'ncov 2019':ti,kw,ab OR 2019nCoV:ti,kw,ab OR 'corono virus':ti,ab,kw OR 'corona virus disease 19':ti,ab,kw OR 'coronavirus 2':ti,ab,kw OR 'covid 19':ti,kw,ab OR 'COVID-19 testing'/exp OR 'SARS-CoV-2 vaccine'/exp OR 'coronavirus receptor'/exp OR ((covid NEAR/2 19):ti,kw,ab) OR ((sars NEAR/2 'cov 2'):ti,kw,ab) OR '2019 ncov':ti,kw,ab OR ((wuhan:ti,kw,ab OR hubei:ti,kw,ab) AND coronavirus*:ti,kw,ab) OR ([2019-2020]/py AND (new:ti,kw,ab OR novel:ti,kw,ab OR pandemic:ti,kw,ab OR epidemic:ti,kw,ab) AND ('coronavirus infection'/exp OR coronavirus*:ti,kw,ab OR coronavirinae/de OR 'corona virus*':ti,kw,ab))) AND 'human'/de AND [embase]/lim NOT ([embase]/lim AND [medline]/lim) AND [english]/lim) NOT ('mental disease'/exp OR depression:ti OR 'systematic review'/exp OR 'review'/exp OR 'editorial'/exp OR 'meta analysis'/exp OR 'case report'/exp OR 'letter'/exp) |
| Cochrane Central | (randomized OR randomly OR randomised OR randomization OR randomisation OR placebo* OR pragmatic OR blind*) AND ("covid 19" OR covid OR covid19 OR "ncov 2019" OR "novel coronavirus" OR "sars cov 2" OR "sars cov-2" OR "ncov 2019" OR sarscov2 OR (wuhan AND coronavirus*) OR "corona virus*" OR "coronavirus disease 2019" OR "coronavirus disease 19" OR "2019 ncov" OR 2019nCoV OR "coronavirus 2") NOT (depression) |

## Footnotes

1 Statistics related to the size COVID-19-related literature were based on the CORD-19 dataset (Lu Wang et al., 2020) and likely represent an underestimation.

